# Modelling and data-based analysis of COVID-19 outbreak in India : a study on impact of social distancing measures

**DOI:** 10.1101/2020.05.12.20099184

**Authors:** Ankit Khushal Barai, Anuradha Singh, Amol Shinde

## Abstract

In this manuscript, we model and visualize the region-wise trends of the evolution to COVID-19 infections employing a SIR epidemiological model. The SIR dynamics are expressed using stochastic differential equations. We first optimize the parameters of the model using RMSE as loss function on the available data using L-BFGS-B gradient descent optimisation to minimise this loss function. This helps to gain better approximation of the model’s parameter for specific country or region. The derived parameters are aggregated to project future trends for the Indian subcontinent for next 180 days, which is currently at an early stage within the infection cycle. The projections are meant to function a suggestion for strategies for the socio-political counter measures to mitigate COVID-19. This study considers the current data for India from various open sources. The SIR models prediction is found following the actual trends till date. The inflection point analysis is important to find out which countries have reached their inflection point of the number of infection.

We found that if current restrictions like lockdown in India continues with same control, then India will observe itś peak in active patients count on 22 May 2020, it will take 28 August 2020 for 90% of the peak active infections to end. Inspired from the study of DDI Lab at Singapore university of technology and design (SUTD), this study additionally tries to model and quantify the variations in the count of active patients in the country which might occur due to effect of waiver in restrictions. It should be noted that these results were predicted using COVID-19 data of India till 03 May 2020.

## 1. Introduction

The novel coronavirus (COVID-19) that has been spreading worldwide since December 2019 has sickened millions of people, shut down major cities, prompted unprecedented global travel restrictions and influenced the global spread of misinformation and panic [1]. COVID-19 has been devastating for the whole world. Some regions are in initial stages of this epidemiology like India. Moreover, India hosts huge population, there is an eminent danger and most of those population are under the poverty-line or the countries do not have huge resources for medical interventions for affected patients. The choices are limited – either choose herd immunity which may backfire and it is too risky or lockdown and restrict the population from traveling. Thus, we attempt to think about the share of population allowed to move around (some are essential services and a few not obeying the orders) and assuming the mobility factors appropriately, we attempt to predict what can happen during the lockdown.

In most of SIR modelling studies [2] done for India, the parameters like *β* and γ were inspired from European nations which do not model the dynamics of India. Some studies [3] make initial guess for parameters by taking *R*(0) = 0. This study deals with the dynamics governing the evolution of the COVID-19 infections are modelled employing a stochastic equation SIR model, inspired from [4] and [5], we study developing model for India specific predictions. We first optimize the parameters of the model using RMSE as loss function on the available data using L-BFGS-B gradient descent optimization to minimise this loss function. This helps to gain better approximation of the model’s parameter for specific country or region. An aggregate of those parameters has been used for projecting the longer-term trends for the Indian region, multiple projections have been generated by varying the exposure factor *E_f_* that influences the growth rate of infections. This is done in order to provide insights for selective quarantining and lockdowns.

The SIR model is one among the only disease models we have to elucidate the spread of an epidemic through a population. Firstly, we explained where the model comes from, including the assumptions that are made and the way the equations are derived, before happening to use the results of the model to answer three important questions:

- will the disease spread?
- what is the utmost number of individuals which will have the disease at one time?
- what percentage people will catch the disease in total?

The answers to these questions are discussed within the context of the present COVID-19 (Coronavirus) outbreak. There is a variation to this model called SIQR model [6] which takes quarantine actions into account.

## 2. Stochastic SIR model

### 2.1. Differential Equations

The differential equations governing the SIR model are as follows:

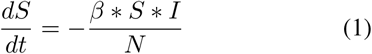

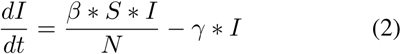

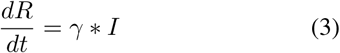

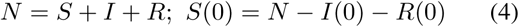

### 2.2. States of model

- I(*t*) is number of active patients at time t.
- *R(t)* is number of recovered+deaths patients at time *t*.
- *t* is the daily-time parameter.
- *dS(t), dI*(*t*), *dR(t)* denotes changes at time *t*.
- *E_f_* denoting the effectiveness of restrictions like lockdown on parameter *β*.

### 2.3. Parameters

- *β* is the factor that models for rise in the number of infections due to interactions between infected and susceptible population.
- *E_f_* denotes the effectiveness of the travel restrictions viz. lockdown etc. A value of *E_f_* < 1 denotes more stricter restrictions than current scenario and *E_f_* > 1 simulates the waiving of the restrictions of social distancing and travel. *E_f_* = 1 models the current restriction scenario.
- *γ* denotes the rate at which infected patients are either getting recovered or died.
- *R*0 is the reproduction factor i.e. the average number of infections caused by an infected host.
- *N* is the approximate to the total population that will get exposed to infection. We have used N = 100,000 which is a good approximation.

## 3. Methodology

### 3.1. Data Analysis

There are a lot of official data sources on the web providing COVID-19 related data. This study makes use of various data sets available on Kaggle.com providing information about COVID-19 in different areas like no. of infections, no. of deaths, testing, age details, week wise data etc. age is one of the important factors in deaths due to infection [7].

For data analysis, various visualisations are created to get better understanding of the various factors affecting the COVID-19 spread. Figure 1, compares the growth of COVID-19 infections in India with other major countries. Similarly, the ratio of recovered:confirmed patients and death:confirmed ratio of India as shown in Figure 2 and Figure 3 clearly indicates the advantage India had over other countries due to immediate steps of lockdown prior to widespread of COVID-19.

**Figure 1.**
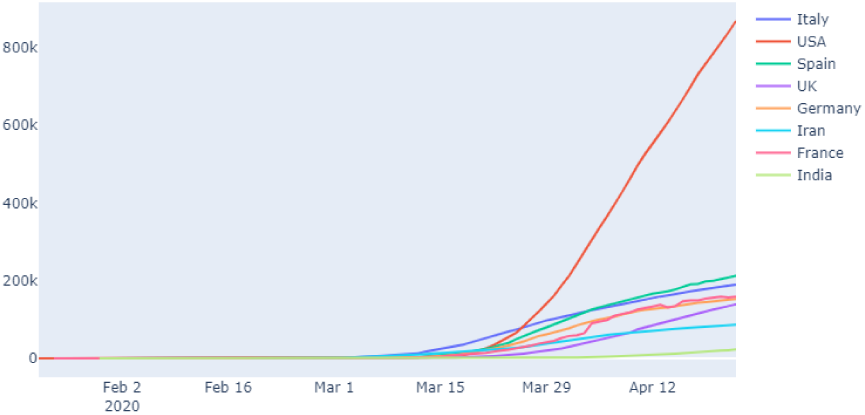
COVID-19 growth in the world

**Figure 2.**
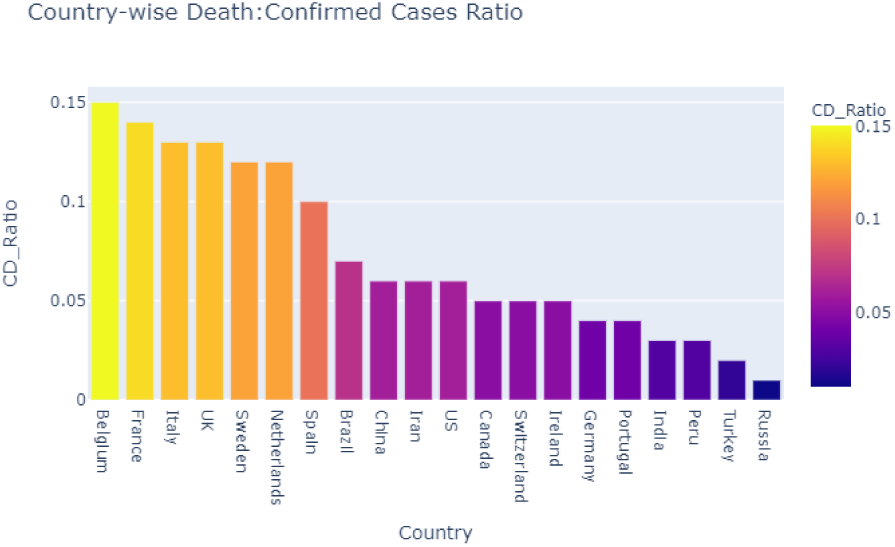
Ratio Death:Confirmed cases

**Figure 3.**
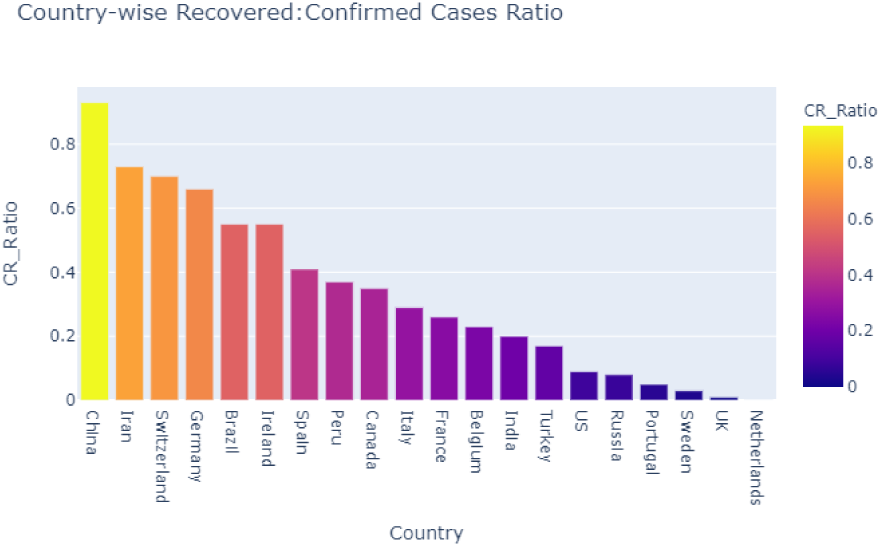
Ratio Recovered:Confirmed cases

It is evident that more than 1 out of 10 person dies in Belgium, France & Italy which is too high. For India, this ratio is nearly one death per 20 confirmed cases. Also, the recovery rate of India is better than countries such as USA and Russia. Age is one of the important factors affecting the number of cases. Figure 4, shows that elderly patients are more prone to deaths related to COVID-19 in India as well as rest of the world. Figure 5, shows that nearly 63.4% of COVID-19 infections are without travel history and Figure 6 shows that probable reasons for such infection spread are contacts with those of travel history, neighbours etc. clearly showing the need of lockdown.

**Figure 4.**
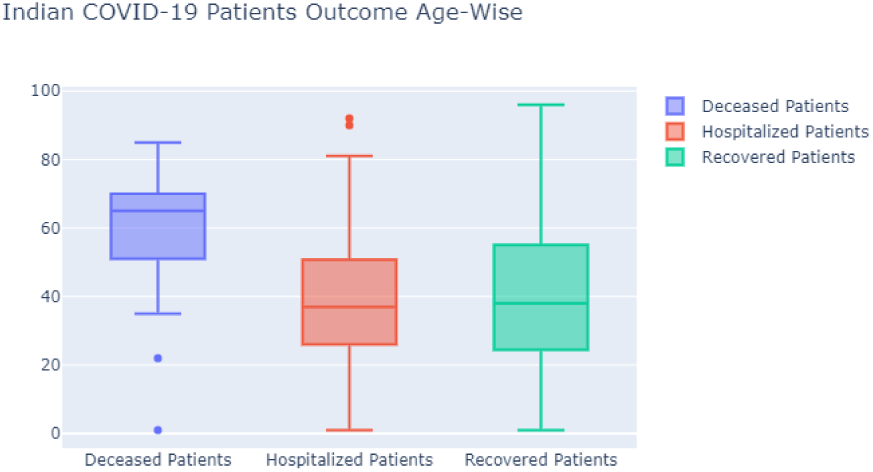
Outcome of cases with respect to age

**Figure 5.**
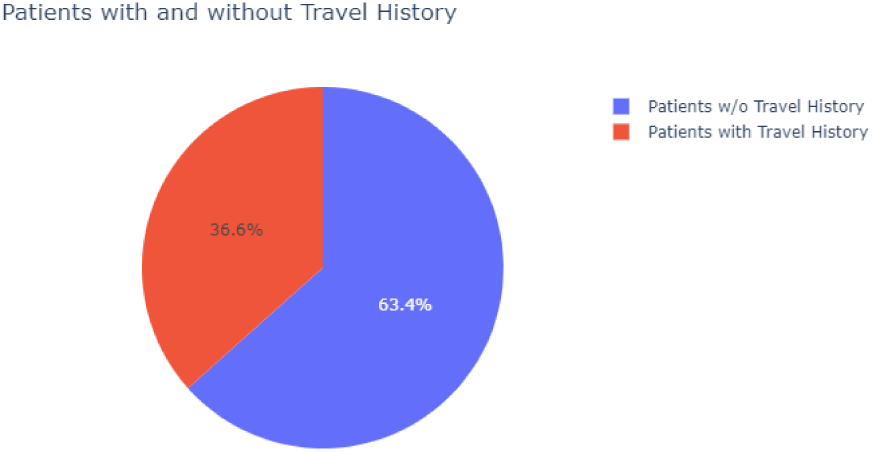
Impact of travel history on COVID-19

**Figure 6.**
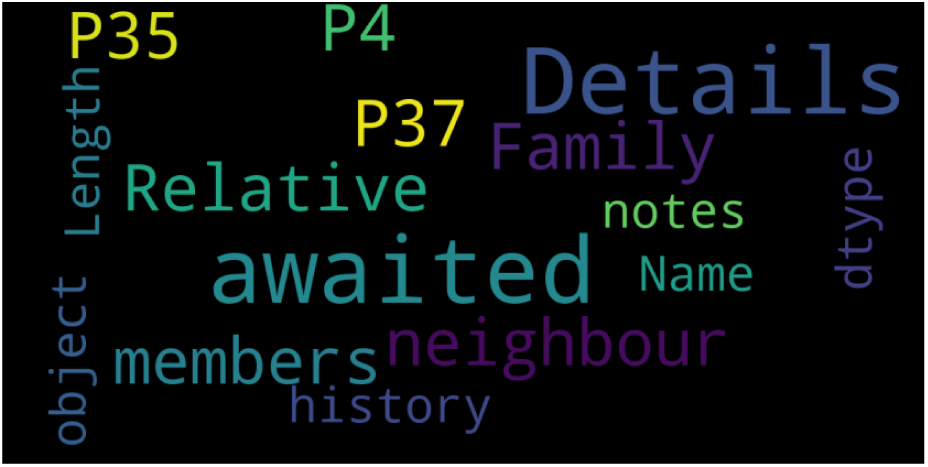
Word cloud for reasons of infection without travel history

### 3.2. SIR modelling analysis

Variation in *E_f_* is used to study the effect of preventive measures like lockdown on the contact rate *β* i.e. effective *β* = *E_f_* * *β* where *E_f_* > 1 means that lockdown is lifted and *E_f_ <* 1 means that lockdown is more stricter than current scenario. On the other hand, *E_f_ >* 1 refers to the situation with more travel permissions than the current lockdown scenarios.

The population *N* which is supposed to have been susceptible is taken as 100,000. The parameters *β* and *γ* are optimized for Indian data by minimizing the RMSE loss on prediction with the existing data available of previous month from 25 March 2020 to 18 April 2020. To simulate the effect of more stricter lockdown and social distancing measures, *E_f_* is used, lower the value of *E_f_*, more stricter the lockdown measures are. By decreasing the *E_f_* i.e. the effectiveness of lockdown is increased and the curve is getting more flattened.

Value of *E_f_* > 1 shows that if lockdown is lifted than curve is shifting towards left showing that more patients will get infected in short time, i.e. the doubling rate of infections becomes smaller, clearly depicting the need of effective social distancing and lockdown. Using algorithm 3 and 3, The predicted curve is found to be following the actual curve for the initial days of prediction and if this model is good approximation, then the model suggests that number of active infected patients will reach peak and start decreasing after 22 May 2020.

### 3.3. Inflection point analysis

The theory behind this is that, when the no. of weekly cases starts dropping consistently for a few consecutive weeks with respect to previous week we have hit the inflection point. From Figure 10 we see that only China and South Korea have been able to hit the inflection point.

Here, we have plotted the log (increased number of confirmed cases with respect to previous week) on *Y* axis and total number of confirmed cases on the *X* axis, till date India’s curve is found to be following similar pattern as rest of the world, only South Korea and China are able to see a dip in this inflection curve.

#### Algorithm 1: SIR algorithm

**Input:** *S, I, R, β, γ, N*

**Output:** 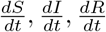

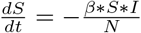

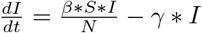

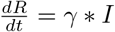

**Table 1.** Variation in reproduction number with *E_f_*

| $\beta = 0.14013338, \gamma = 0.0180899$ and Effective $\beta = \beta * E_f$ | |
| --- | --- |
| $E_f$ | Reproduction Number $R_0$ |
| 0.25 | 1.24 |
| 0.5 | 2.49 |
| 0.75 | 3.74 |
| 1.00 | 4.98 |
| 1.25 | 6.23 |
| 1.50 | 7.48 |
| 1.75 | 8.73 |

#### Algorithm 2: Model Prediction

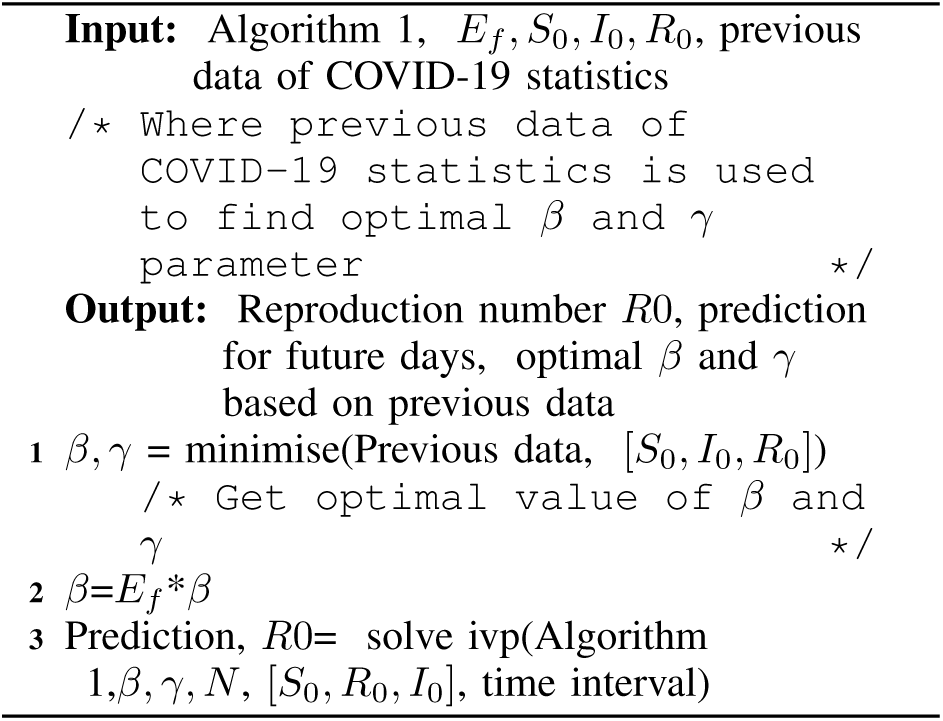

## 4. Results and Discussions

Table1 enlists the reproduction number *R*0 obtained for different values of *E_f_* showing that if lockdown is not implemented strictly, the reproduction number will get increased creating large burden on the healthcare system of the country.

From Figure 7, it is evident that the curve shifts to left with increase in *E_f_*, which means that when restrictions are waived off, the number of infections will be high and growth rate will also be very high. If current restrictions are continued i.e. *E_f_* = 1, then total number of active infections will start to decrease after 22 May 2020. These results are in accordance with the study by DDI lab, Singapore university of technology and design [8]. In addition, this study also models the effect of any waivers on social restrictions with *E_f_* > 1, the total number of infections are expected to increase before reaching its peak. From Figure 7 and 8, with waiver in restrictions, the number of new infections will be much higher than the recovery. Figure 9 shows that with help of restrictions we can control the number of infections as well as the rate at which are approaching the peak of curve.

**Figure 7.**
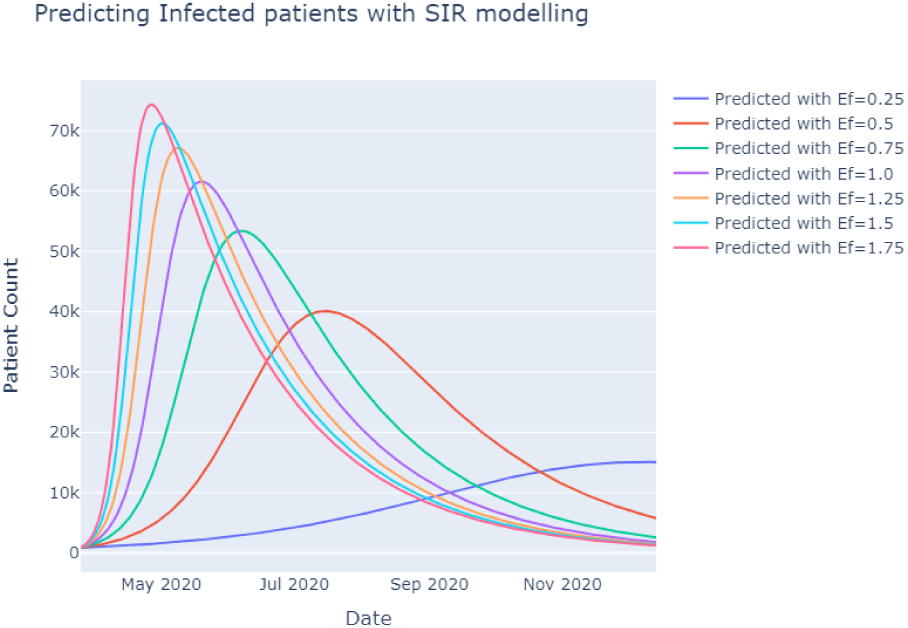
SIR modelling infected patients prediction

**Figure 8.**
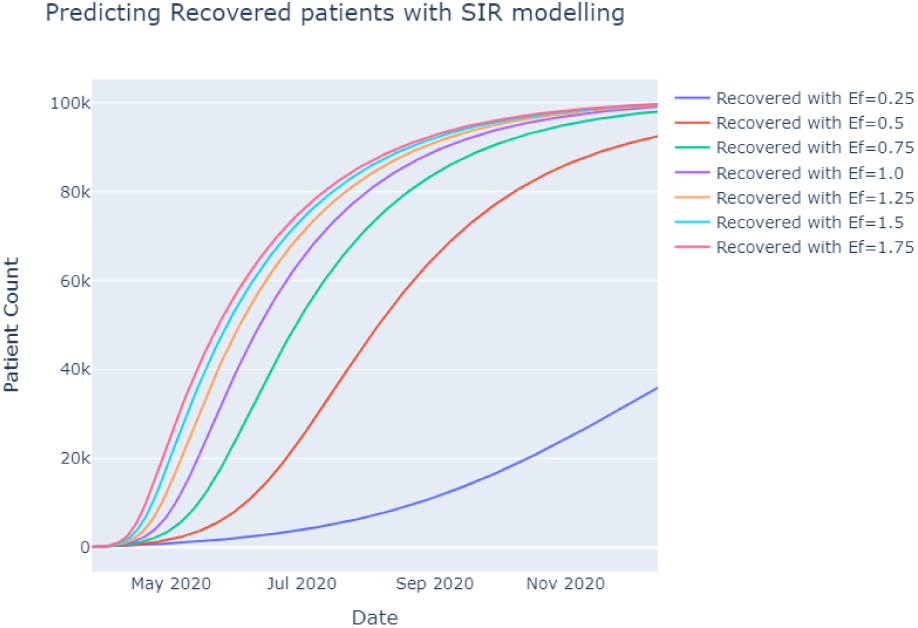
SIR modelling recovered patients prediction

**Figure 9.**
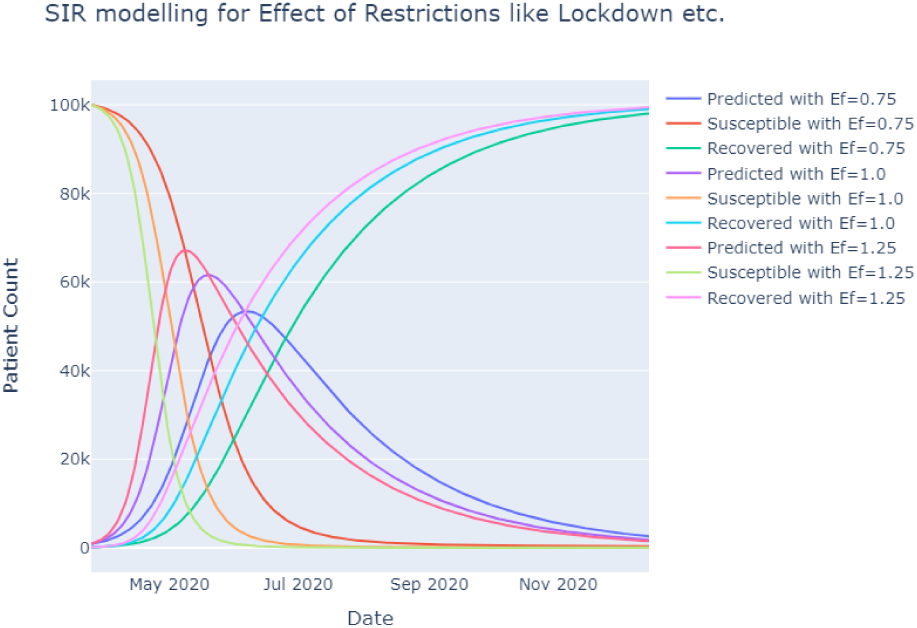
SIR modelling with restriction impact factor *E_f_*

From Figure 10, it is evident that only China and South Korea have reached the inflection point which means that infections are yet to reach it’s peak in India. The model tells us that to scale back the impact of the disease we would like to lower the ‘contact ratio’ the maximum amount as possible - which is strictly what the present social distancing measures are designed to try to do. Figure 11, for *E_f_* = 1, it shows that the count of daily new cases per day will decrease to 97% of its peak on 16 July 2020 and 99% on 8 August 2020.

**Figure 10.**
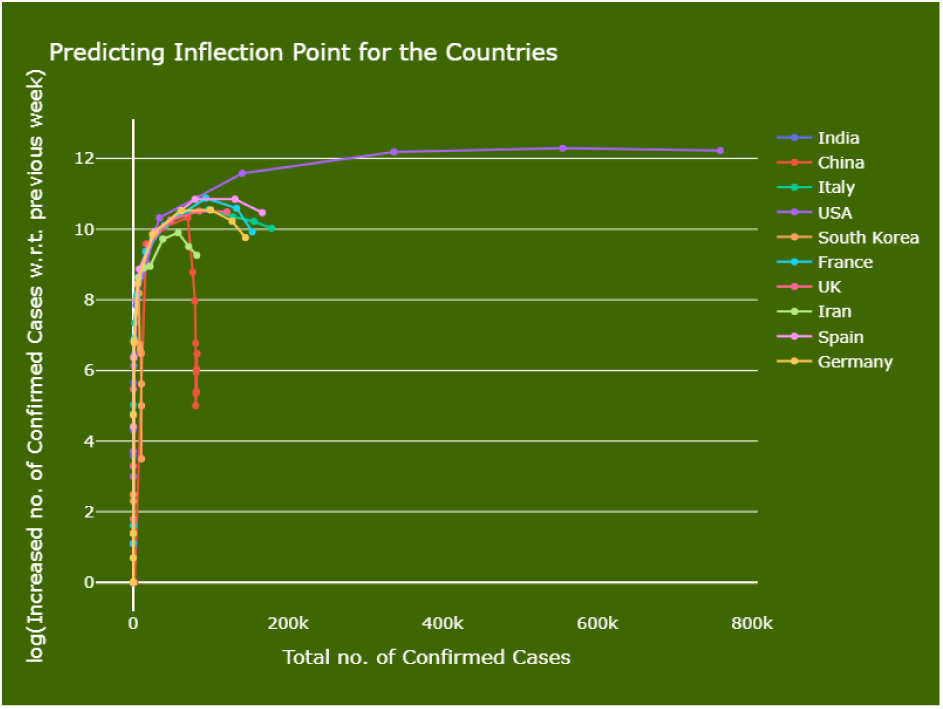
Country-wise inflection point analysis on COVID-19 data

**Figure 11.**
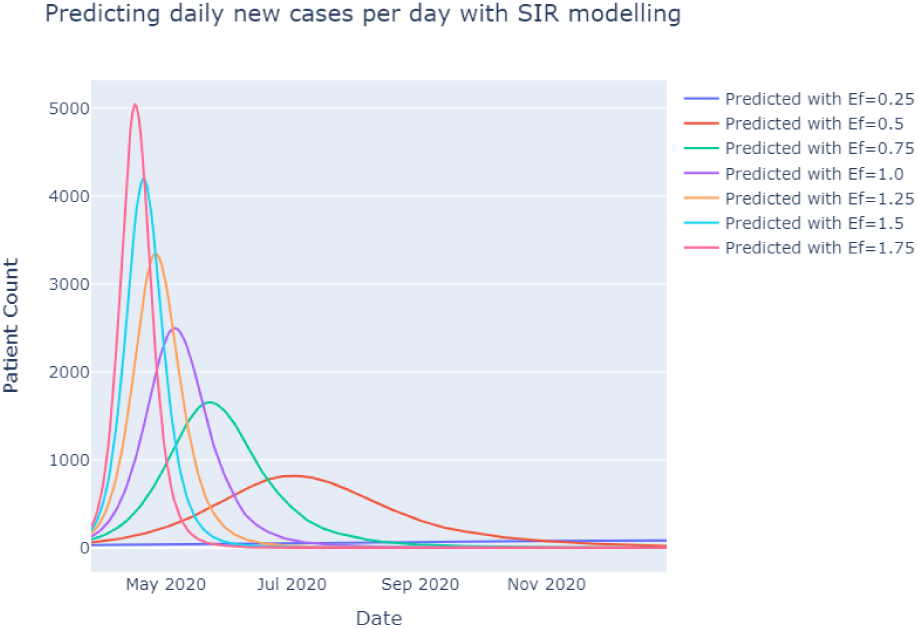
Daily new cases/day predicted by SIR model

## 5. Conclusion

Only China and South Korea are able to reach their inflection points as of now. We found that if current restrictions like lockdown in India continues with same control till 22 May 2020 then, the number of active infected patients will achieve its peak and start decreasing after this date. To model the impact of travel restrictions like lockdown, social distancing on the reproduction number; we used a parameter *E_f_* whose value is used to simulate the effectiveness of social restrictions.

With more travel restriction like lockdown the value of *E_f_* decreases and hence, effective contact rate decreases which results in reduced *R*0 [9]. Reproduction number is the average number of non-infected people who got infected by a infected patient. By practising social distancing and lockdown the value of *E_f_* decreases. Any waiver in the current restriction will result in higher reproduction number *R*0 as well as increase in number of infections in very short time. In addition to lockdown, rapid testing strategies are required [10] for the faster infection detection and thus helping in lifting up lockdown and revival of economy.

## Data Availability

https://www.kaggle.com/sudalairajkumar/novel-corona-virus-2019-dataset
• https://www.kaggle.com/parulpandey/coronavirus-cases-itesting-rate-all-countries
• https://www.kaggle.com/sudalairajkumar/covid19-in-india

